# Cross-informant validation of the Pica, ARFID, and Rumination Disorder Interview in adolescents: Agreement between child and parent reports

**DOI:** 10.64898/2026.11.16.26367277

**Authors:** Felicia Dahlbäck, Tom Moberg, Diana Friskson, Laila L. Myrberg, Liv Hog, Nadia Micali, Cynthia M. Bulik, Lisa Dinkler

**Author notes:** Correspondence should be addressed to Lisa Dinkler, PhD, Department of Medical Epidemiology and Biostatistics, Karolinska Institutet, Nobels väg 12a, 17165 Solna, Sweden. Author details: Diana Friskson, Tom Moberg, Felicia Dahlbäck, Laila L. Myrberg, Liv Hog, MSc, Nadia Micali, MD, PhD, Cynthia M. Bulik, PhD.

## Abstract

**Background:** Reliable assessment of avoidant/restrictive food intake disorder (ARFID) in young people may require information from both children and caregivers, yet cross-informant agreement has not been examined for the Pica, ARFID, and Rumination Disorder Interview (PARDI). This study evaluated agreement between child and parent PARDI interviews for ARFID diagnosis, individual DSM-5 Criterion A, ARFID dimensions, and overall severity.

**Methods:** Twenty children aged 11–14 years screening positive for ARFID and one parent per child were recruited from the ARFID InitiativE Sweden (ARIES) cohort. Children and parents completed separate PARDI interviews with trained interviewers. Parent–child agreement was assessed using percentage agreement and Cohen’s kappa (κ) for categorical ratings of ARFID diagnosis and Criterion A consequences. For continuous ratings of ARFID dimensions and overall severity, agreement was assessed using intraclass correlation coefficients (ICCs) and paired t-tests comparing mean parent and child scores.

**Results:** Parents gave higher mean scores than children across all continuous domains, although the difference was statistically significant only for overall severity (raw mean difference=0.42, p=.003). Parent–child agreement was poor for sensory sensitivity (ICC=0.42), moderate for lack of interest (ICC=0.63) and overall severity (ICC=0.71), and excellent for concern about aversive consequences (ICC=0.95). ARFID was identified in 17/20 parent interviews and 14/20 child interviews; diagnostic agreement was 75% (κ=0.31, p=.133). Agreement was perfect (100%) for Criterion A1, high for A2 and A3 (both 95%), and lowest for psychosocial impairment (A4; 65% agreement, κ=0.19). Item-level findings indicated a general tendency toward higher parental ratings, although children provided higher ratings on selected items involving food restriction, sensory experiences, hunger cues, and abdominal pain.

**Conclusions:** Child and parent PARDI interviews provided overlapping but non-interchangeable information. Agreement varied across different aspects of ARFID, with greater discrepancies for psychosocial impairment and sensory sensitivity. These differences may partly reflect what children and parents are able to observe or experience directly. Multi-informant assessment may therefore offer a more complete account of pediatric ARFID than either response alone.

**Plain English Summary:** Children may experience and describe avoidant/restrictive food intake disorder (ARFID) symptoms differently from how their parents observe and report them. We compared separate child and parent diagnostic interviews using the Pica, ARFID, and Rumination Disorder Interview (PARDI) in 20 pairs to understand how closely their assessments of ARFID agreed. Parents and children often provided similar information, but agreement differed depending on what was being assessed. They agreed more closely about effects on weight, growth, and nutrition, and about fears of aversive consequences of eating such as choking or vomiting. Agreement was lower for how eating difficulties affected daily and social life, for lack of interest in food or eating, and for sensitivity to the taste, texture, or smell of food. Parents generally rated the overall severity of eating difficulties higher than children, although children gave higher ratings for some experiences, such as difficulty recognizing hunger, discomfort with messy or sticky foods, and fear of abdominal pain. These findings suggest that children and parents provide overlapping but not identical information about ARFID. Child and parent accounts may each highlight different aspects of ARFID, making both perspectives valuable sources of information during assessment.

## Background

Avoidant/restrictive food intake disorder (ARFID) is a feeding disorder characterized by persistent avoidance or restriction of the amount and/or variety of food intake without weight- or shape-related psychopathology that defines several other eating disorders (1). The eating disturbance is typically driven by one or more partially overlapping dimensions: sensory sensitivity to food characteristics (e.g., smell, taste, and texture), lack of interest in food or eating, and concern about aversive consequences such as choking, vomiting, or abdominal pain (1, 2). Population-based studies estimate the prevalence of ARFID at approximately 1–2%, with broadly similar rates across sexes (3–5). Although onset can occur across the lifespan, ARFID commonly begins in childhood (6, 7). Therefore, assessments likely occur in a pediatric context, where multi-informant assessment is the norm. The Pica, ARFID, and Rumination Disorder Interview (PARDI) is a semi-structured clinical interview designed to assess the presence and severity of ARFID and includes separate child and parent versions to support cross-informant assessments (8). Understanding the extent to which child and parent informants agree is particularly important when only one perspective is available, as differences between informants may affect diagnostic classification and estimates of symptom severity. The present study addresses this need by evaluating PARDI cross-informant agreement for ARFID diagnosis, individual diagnostic criteria, ARFID dimensions, and overall severity.

To meet diagnostic criteria for ARFID, restricted food intake must result in at least one clinically significant consequence: weight loss or faltering growth (Criterion A1), nutritional deficiency (A2), dependence on enteral feeding or oral nutritional supplements (A3), or marked interference with psychosocial functioning (A4). These consequences differ in how directly they can be observed, making informant perspective particularly relevant in pediatric assessment. Whereas children have privileged access to internal experiences such as disgust, anxiety, appetite, and interoceptive cues, parents are often better positioned to observe intake, growth, supplement use, and impairment across daily routines. Criterion A4 (psychosocial impairment) may be particularly susceptible to parent–child disagreement, as different aspects of impairment are likely to vary in salience between informants. Children may be more attuned to the immediate subjective impact of their eating difficulties, such as embarrassment and anxiety when eating with peers—experiences that may not be readily apparent to parents. Parents, in turn, may be better positioned to recognize broader patterns of functional impairment, such as reduced participation in everyday activities, disruption of routines, or the need for ongoing accommodation around eating.

ARFID cross-informant validation studies are still rare, but the few available studies suggest that parent–child reports are related but not interchangeable (9, 10). In a caregiver-youth study using the Nine Item ARFID Screen (NIAS; 11), caregivers reported higher scores than children across all three subscales, especially for low appetite, and caregiver ratings more often crossed screening thresholds (9). Another study comparing parental and adolescent ratings found no significant differences on any of the NIAS subscales, although some differences emerged at the item level (12). Using the Eating Disorder Examination (EDE)-ARFID module, Schmidt et al. (10) found high parent-child agreement for diagnosis while agreement varied across individual diagnostic criteria. Parent–child discordance is not specific to ARFID, with previous studies demonstrating informant differences in child and adolescent eating disorder assessments. Swanson et al. (13), for example, found low concordance between parental and adolescent reports of eating disorder symptoms at ages 14 and 16, with patterns of discordance varying across symptoms. Mariano et al. (14) found that children younger than 13 years reported less severe cognitive eating-disorder symptoms than their parents, whereas adolescents aged 13–18 years generally reported greater symptom severity than their parents. Together, these findings suggest that parent–child agreement can vary depending on the symptoms being assessed, what each informant is able to observe or report, and the child’s developmental stage. Patterns of agreement and disagreement can therefore reveal how eating difficulties are captured from child and parent perspectives. The clinical relevance of incorporating multiple perspectives is supported by House et al. (15), who found that a cross-informant assessment showed closer agreement with multidisciplinary clinical ED diagnosis than an individual interview with the young person alone.

ARFID assessment instruments, including the PARDI, continue to undergo validation (16). Despite the PARDI’s explicit cross-informant design, parent–child agreement for its diagnostic and dimensional ratings has not previously been reported. The present study addresses this gap by comparing parent–child dyad PARDI interviews in 11–14-year-old children screening positive for ARFID in the ARFID InitiativE Sweden (ARIES) cohort. We examined agreement for ARFID diagnosis, diagnostic criteria A1–A4, the three ARFID dimensions, overall severity, and individual PARDI items. Mapping both convergence and disagreement between parent and child ratings may help determine which aspects of ARFID are consistently captured across informants, when combining perspectives provides clinically useful additional information, and may help inform effective treatment planning.

## Method

### Study design and participants

Participants were recruited from ARFID InitiativE Sweden (ARIES), a Swedish cohort study investigating genetic and environmental factors associated with ARFID in children aged 6-14 years (17). Participants were recruited via social media, healthcare services, school health services, and advocacy organizations. Enrollment occurred between May 2024 and February 2026. Parents completed online screening questionnaires and additional measures concerning eating behavior, health, neurodevelopmental and psychiatric conditions, quality of life, and family characteristics.

ARIES screening cases were required to (1) screen positive for ARFID on either the parent-report version of PARDI–ARFID Questionnaire (PARDI-AR-Q; 18) or the parent-report version of NIAS, (2) screen negative for clinically significant non-ARFID eating disorder symptoms on the Parent Eating Disorder Examination Questionnaire (PEDE-Q; 19), and have no lifetime diagnosis of anorexia nervosa or bulimia nervosa (Table 1). This approach has demonstrated good diagnostic performance in accurately identifying probable ARFID and excluding non-cases when evaluated against the PARDI (20).

**Table 1.**
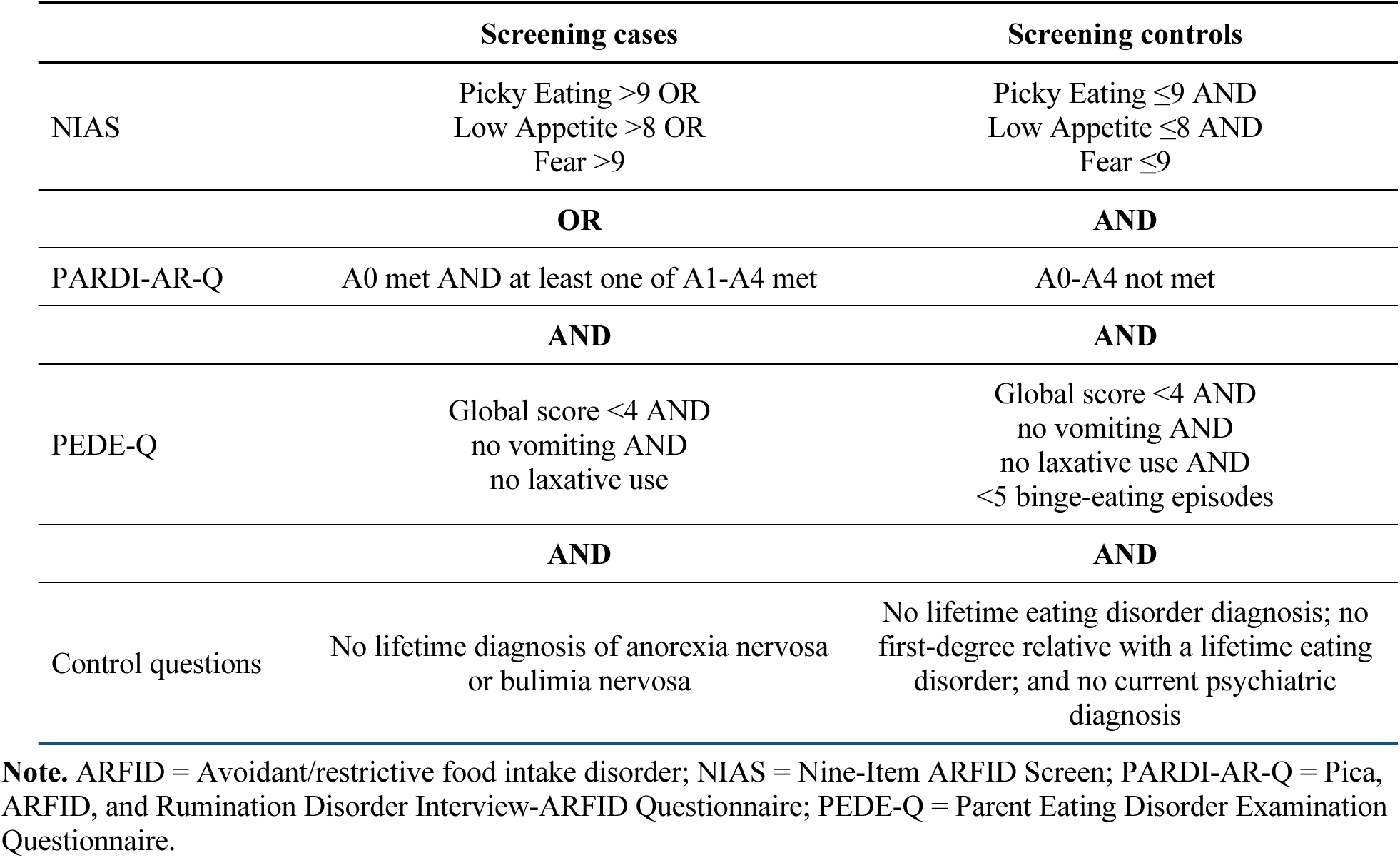
ARIES screening criteria for cases and controls (adapted from Hog et al., 2025).

|  | **Screening cases** | **Screening controls** |
|---|---|---|
| NIAS | Picky Eating >9 OR<br>Low Appetite >8 OR<br>Fear >9 | Picky Eating ≤9 AND<br>Low Appetite ≤8 AND<br>Fear ≤9 |
|  | **OR** | **AND** |
| PARDI-AR-Q | A0 met AND at least one of A1-A4 met | A0-A4 not met |
|  | **AND** | **AND** |
| PEDE-Q | Global score <4 AND<br>no vomiting AND<br>no laxative use | Global score <4 AND<br>no vomiting AND<br>no laxative use AND<br><5 binge-eating episodes |
|  | **AND** | **AND** |
| Control questions | No lifetime diagnosis of anorexia nervosa or bulimia nervosa | No lifetime eating disorder diagnosis; no first-degree relative with a lifetime eating disorder; and no current psychiatric diagnosis |

**Note.** ARFID = Avoidant/restrictive food intake disorder; NIAS = Nine-Item ARFID Screen; PARDI-AR-Q = Pica, ARFID, and Rumination Disorder Interview-ARFID Questionnaire; PEDE-Q = Parent Eating Disorder Examination Questionnaire.

For the present study, children aged 11–14 years who had been classified as ARIES screening cases were eligible. At the time of sampling, 451 children in this age range met the screening case requirement. To ensure that screening and interview assessments reflected a similar time period, invitations were sent no later than seven weeks after parents had consented to participation in ARIES. Sixty-six parent-child pairs were invited, with invitations stratified by sex to obtain a broadly balanced distribution (37 boys and 29 girls). Twenty-nine pairs (44%) initially agreed to participate. Four children withdrew after their parent had been interviewed, two pairs were excluded because the interviews could not be completed as required, two pairs cancelled before interview, and one pair did not respond to scheduling attempts. Twenty pairs (69%) completed both interviews and constituted the analytic sample (Figure 1).

**Figure 1.**
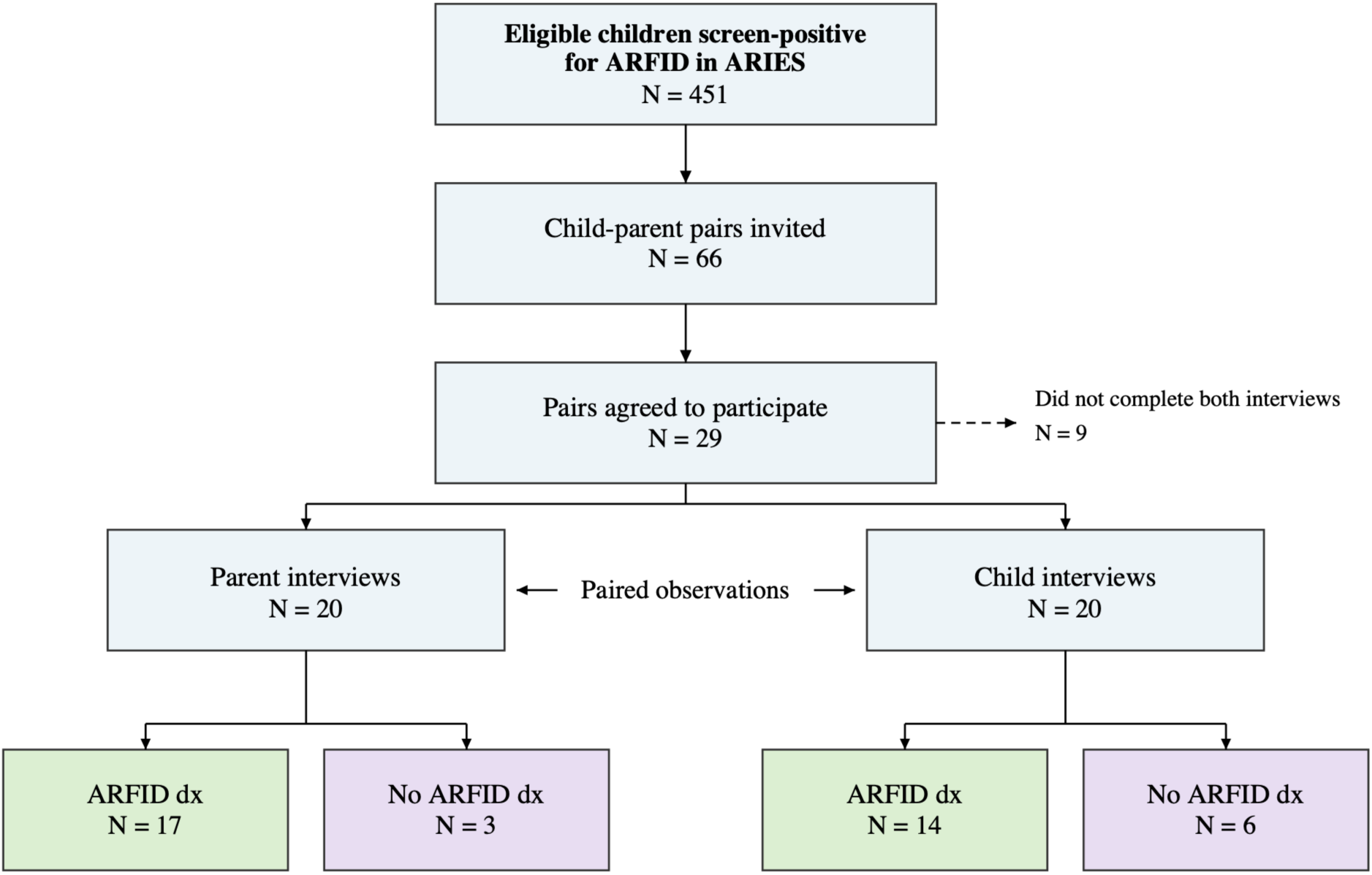
Recruitment, participation, and diagnostic outcomes for the 20 child-parent pairs. Of the nine pairs who did not complete both interviews, four children withdrew after the parent interview, two pairs could not complete the interviews as required, two pairs cancelled before interview, and one pair did not respond to scheduling. ARFID = avoidant/restrictive food intake disorder; ARIES = ARFID InitiativE Sweden; PARDI = Pica, ARFID, and Rumination Disorder Interview.

### Measures

The Pica, ARFID, and Rumination Disorder Interview (PARDI) is a semi-structured, cross-informant interview developed to assess pica, ARFID, and rumination disorder (8, 21). Age-appropriate child and parent versions include screening for competing eating disorders and relevant medical or psychiatric conditions, followed by diagnostic and dimensional assessments. Items are answered dichotomously or rated on a 0–6 scale. The ARFID section yields dimensional mean scores for sensory sensitivity, lack of interest in food or eating, and concern about aversive consequences, together with an overall severity score, where higher scores indicate greater symptom severity.

Based on DSM-5 criteria, an ARFID diagnosis requires (1) presence of avoidant or restrictive eating, (2) presence of at least one consequence under Criteria A1-A4, (3) duration exceeding one month, and (4) fulfillment of exclusion requirements: the eating disturbance must not be better explained by (1) lack of available food or a culturally sanctioned practice (Criterion B) or (2) anorexia nervosa, bulimia nervosa, or related weight- or shape psychopathology (Criterion C), and, when a medical, neurodevelopmental, or other mental condition is present, the eating disturbance exceeds what would ordinarily be expected from that condition and warrants additional clinical attention (Criterion D).

### Procedure

Child and parent interviews were conducted separately by video conference between April and November 2025 and were recorded for quality assurance. The interviewer group comprised one doctoral student, two medical students, and two undergraduate psychology students. All interviewers completed six hours of training that included an introduction to ARFID and the PARDI, independent rating of a recorded interview, and group discussion of the training interview ratings. After the training, all interviewers completed a supervised interview that was subsequently reviewed with the study team. Difficult ratings were reviewed in bi-weekly consensus meetings throughout data collection. Different interviewers assessed the child and parent in each pair to reduce direct carryover of information. Parent interviewers were blinded to ARIES screening status and did not have access to questionnaire data during interview administration or scoring. As only children who screened positive for ARFID were interviewed, child interviewers were aware of their screening status. Current height and weight of the child were reported by both the parent and the child, and parent–child reported values were highly correlated (r = .99) for both height and weight. Sex- and age-adjusted standard deviation scores (SDS) were calculated for parent-reported height, weight, and body mass index (BMI) using Swedish age- and sex-specific reference norms (22, 23). Pairs where only one member could provide current anthropometric information (i.e., either the child or the parent; n=6) were excluded from analyses specific to Criterion A1.

### Statistical analysis

Analyses were performed in R version 4.5.0, with statistical significance set at p < .05. Cross-informant agreement for ARFID diagnosis and Criteria A1-A4 (categorical ratings) was quantified using percentage agreement and Cohen’s kappa (κ). κ values were interpreted as slight (0.00–0.20), fair (0.21–0.40), moderate (0.41–0.60), substantial (0.61–0.80), or almost perfect (0.81–1.00) (24). McNemar’s tests assessed whether parent–child disagreements showed a systematic directional pattern, such as parents endorsing a criterion more often than children when the two informants disagreed (25). Intraclass correlation coefficients (ICCs) with 95% confidence intervals were used for sensory sensitivity, lack of interest, concern, and overall severity (continuous ratings). ICCs below 0.50 were interpreted as poor, 0.50–0.75 as moderate, 0.75–0.90 as good, and above 0.90 as excellent (26). Paired t-tests examined mean differences between parent and child scores. At the item level, mean parent-minus-child differences assessed the direction and magnitude of rating discrepancies, with positive values indicating higher parental ratings and negative values higher child ratings. Spearman’s rho assessed rank-order correspondence, that is, the degree to which higher parental ratings were associated with higher child ratings across dyads (27). Interrater reliability was evaluated with Cohen’s kappa for categorical ratings and ICCs for continuous scores. There were no missing data, as all PARDI diagnostic ratings were completed during the interview.

### Interrater reliability of interviewers

During the initial phase of data collection, 23 interviews (17 parent and six child interviews) were independently double rated to evaluate interviewer reliability. Interrater reliability estimates were calculated using interviewers’ original independent ratings prior to consensus discussions. Diagnostic classifications used in subsequent analyses were based on consensus ratings after discrepant cases had been reviewed and resolved. Reliability between interviewers was moderate to excellent. Agreement for ARFID diagnosis was 91.3% (κ = 0.80, p < .0001). Agreement was 82.6% for Criterion A1 (κ=0.55, p=0.0085), 100% for Criterion A2 (κ=1.00, p<0.0001), 95.7% for Criterion A3 (κ=0.83, p<0.0001), and 82.6% for Criterion A4 (κ=0.62, p=0.0031). Agreement for the exclusion Criteria B–D was 100%. ICCs indicated excellent reliability for Sensory Sensitivity (ICC=0.89, 95% CI [0.77, 0.95]), Lack of Interest (ICC=0.94, 95% CI [0.87, 0.98]), and Severity (ICC=0.96, 95% CI [0.92, 0.98]), and good reliability for Concern of aversive consequences (ICC=0.77, 95% CI [0.54, 0.90]).

### Ethical approval

The study was approved by the Swedish Ethical Review Authority (approval nos. 2023-04638-0 and 2025-01356-02). Parents received written study information and provided written consent in the ARIES study and oral consent in the sub-study including PARDI interviews. Children received age-adapted information emphasizing that participation was voluntary and provided oral assent before the interview. Recordings were stored on a password-protected SharePoint site accessible only to the research team and are scheduled for deletion six months after study completion.

## Results

### Participant characteristics

The final sample comprised 20 parent-child pairs (13 girls and 7 boys). Given the small sample, no sex-stratified analyses were undertaken. Mean age at the parent interview was 13.4 years (range 11.6–14.9). On average, the parent interview occurred seven days after the child interview. Parent-reported mean SDS for weight, height, and BMI were -0.6, 0.1, and -0.9, respectively (Table 2). For 18 of the 20 children, the same parent completed the ARIES screening and the PARDI interview. For one child, a different parent participated in the PARDI interview, and for another, both parents had completed the ARIES screening together, although only one was interviewed. Nineteen parent interviews were completed by the mother and one by the father. Parent-reported co-occurring conditions included attention-deficit/hyperactivity disorder (n = 7), constipation (n = 5), autism (n = 3), asthma (n = 3), and anxiety disorders (n = 3).

**Table 2.** Participant characteristics.

| **Characteristic** | **N = 20** |
|---|---|
| Sex |  |
| Female | 13 (65%) |
| Male | 7 (35%) |
| Age at parent interview, years | 13.4 (1.0); 11.6–14.9 |
| Days between ARIES consent and parent interview | 42.5 (17.8); 13.5–77.6 |
| Days between ARIES consent and child interview | 49.1 (16.9); 21.8–79.1 |
| Weight-SDS (parent-reported) | -0.6 (1.1); -3.0–1.7 |
| Height-SDS (parent-reported) | 0.1 (1.0); -1.4–2.5 |
| BMI-SDS (parent-reported) | -0.9 (1.2); -3.0–1.2 |

**Note.** Categorical variables are n (%). Continuous variables are mean (SD); range. BMI = body mass index; SD = standard deviation.

### Agreement for ARFID dimensions and overall severity

Parents’ mean scores for all three ARFID dimensions and overall severity were higher than children’s, however only the difference in overall severity was statistically significant (ΔM = 0.42, p = .003; Table 3). Mean parent-minus-child differences were 0.20 for sensory sensitivity, 0.33 for lack of interest, and 0.09 for concern (all p > 0.05). Cross-informant agreement differed markedly by domain. Agreement was poor for sensory sensitivity (ICC = 0.42, 95% CI [-0.02, 0.72]), moderate for lack of interest (ICC = 0.63, 95% CI [0.29, 0.83]), excellent for concern (ICC = 0.95, 95% CI [0.87, 0.98]), and moderate for overall severity (ICC = 0.71, 95% CI [0.25, 0.89]; Table 3 & Figure 2). The wide confidence intervals for sensory sensitivity, lack of interest, and overall severity indicated substantial uncertainty around the point estimates.

**Figure 2.**
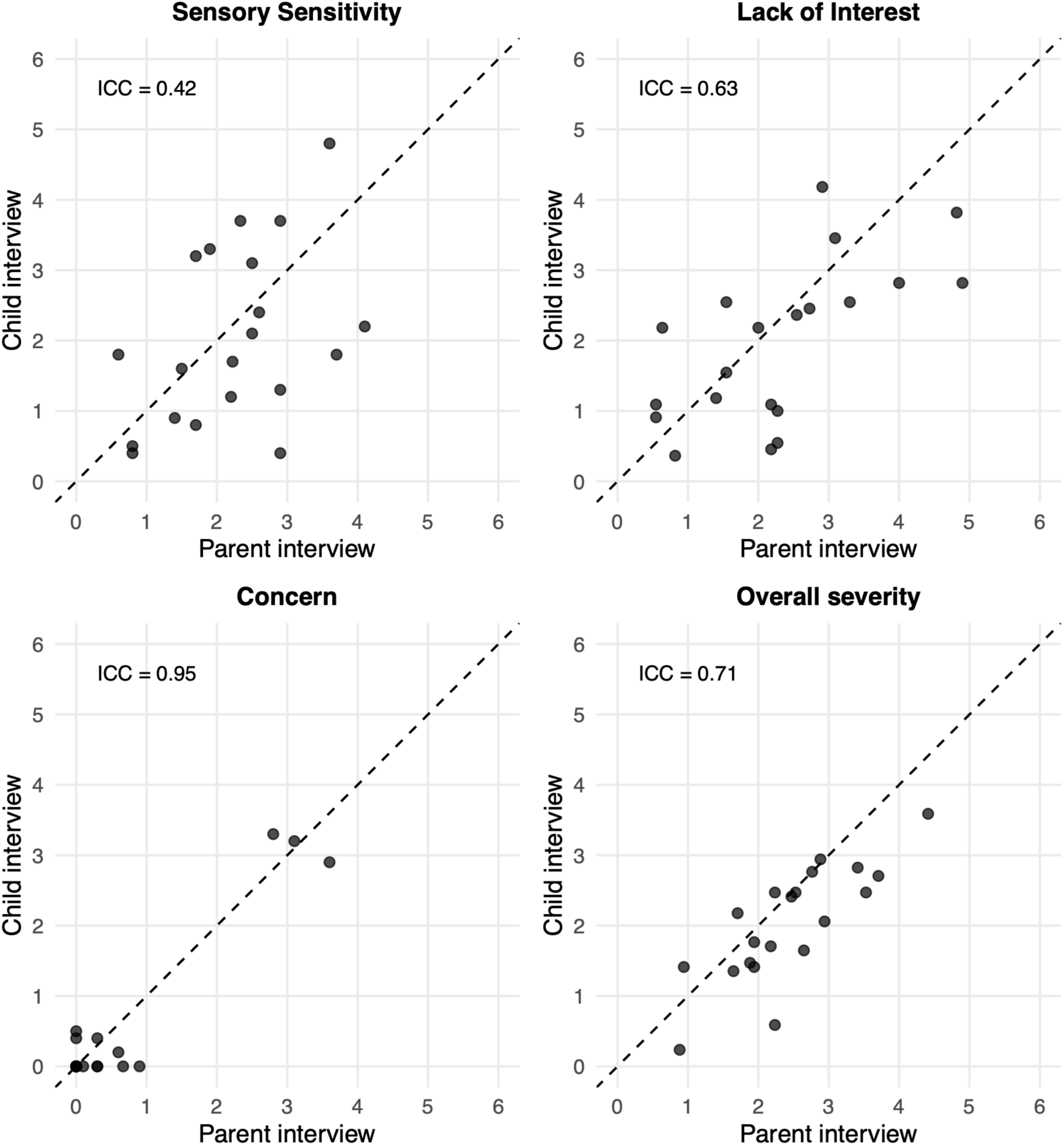
Scatterplots of parent and child ratings for the three ARFID dimensions and overall severity. Dashed diagonal represents identical parent and child scores; panel labels report intraclass correlation coefficients (ICC). ARFID = Avoidant/restrictive food intake disorder; PARDI = Pica, ARFID, and Rumination Disorder Interview.

**Table 3.** Parent-child agreement on PARDI dimensions and overall severity (continuous ratings).

|  |  |  | Mean differences |  |  | Agreement |  |  |
|---|---|---|---|---|---|---|---|---|
| **Domain** | **Parent<br>M (SD),<br>Range** | **Child<br>M (SD),<br>Range** | **ΔM (SD)** | **t<br>(paired<br>t-test)** | **p** | **ICC** | **95% CI** | **p** |
| Sensory sensitivity | 2.24 (0.97),<br>0.60–4.10 | 2.04 (1.25),<br>0.60–4.10 | 0.20 (1.21) | 0.73 | .475 | 0.42 | -0.02–0.72 | .031 |
| Lack of interest | 2.31 (1.29),<br>0.55–4.90 | 1.98 (1.13),<br>0.55–4.90 | 0.33 (1.02) | 1.47 | .159 | 0.63 | 0.29–0.83 | .001 |
| Concern | 0.63 (1.13),<br>0.00–3.60 | 0.55 (1.13),<br>0.00–3.60 | 0.09 (0.37) | 1.06 | .301 | 0.95 | 0.87–0.98 | .001 |
| Overall severity | 2.44 (0.89),<br>0.88–4.41 | 2.02 (0.82),<br>0.88–4.41 | 0.42 (0.55) | 3.40 | .003 | 0.71 | 0.25–0.89 | .003 |

**Note.** ΔM = Parent score – Child score. ICC = Intraclass correlation coefficient; CI = Confidence interval.

### Agreement for ARFID diagnosis and individual diagnostic criteria

ARFID diagnostic criteria were met in 17 parent interviews (85.0%) and 14 child interviews (70.0%). Informants agreed on an ARFID diagnosis in 13 pairs (65.0%) and on no diagnosis in two pairs (10.0%); five pairs were discordant (25.0%; Figure 3). Thus, overall diagnostic agreement was 75.0%, with fair but non-significant agreement beyond chance (κ = 0.31, p = .133; Table 4). Among the five discordant pairs, four were classified as ARFID based on the parent interview but not the child interview, whereas one pair showed the opposite pattern. All five diagnostic discrepancies corresponded to disagreement on Criterion A4 (psychosocial impairment): A4 was met in the parent but not the child interview in the four parent-positive/child-negative pairs, and in the child but not the parent interview in the remaining pair. This directional imbalance was not statistically significant (McNemar’s χ² = 0.80, p = .371). Age distributions were broadly similar between concordant and discordant pairs.

**Figure 3.**
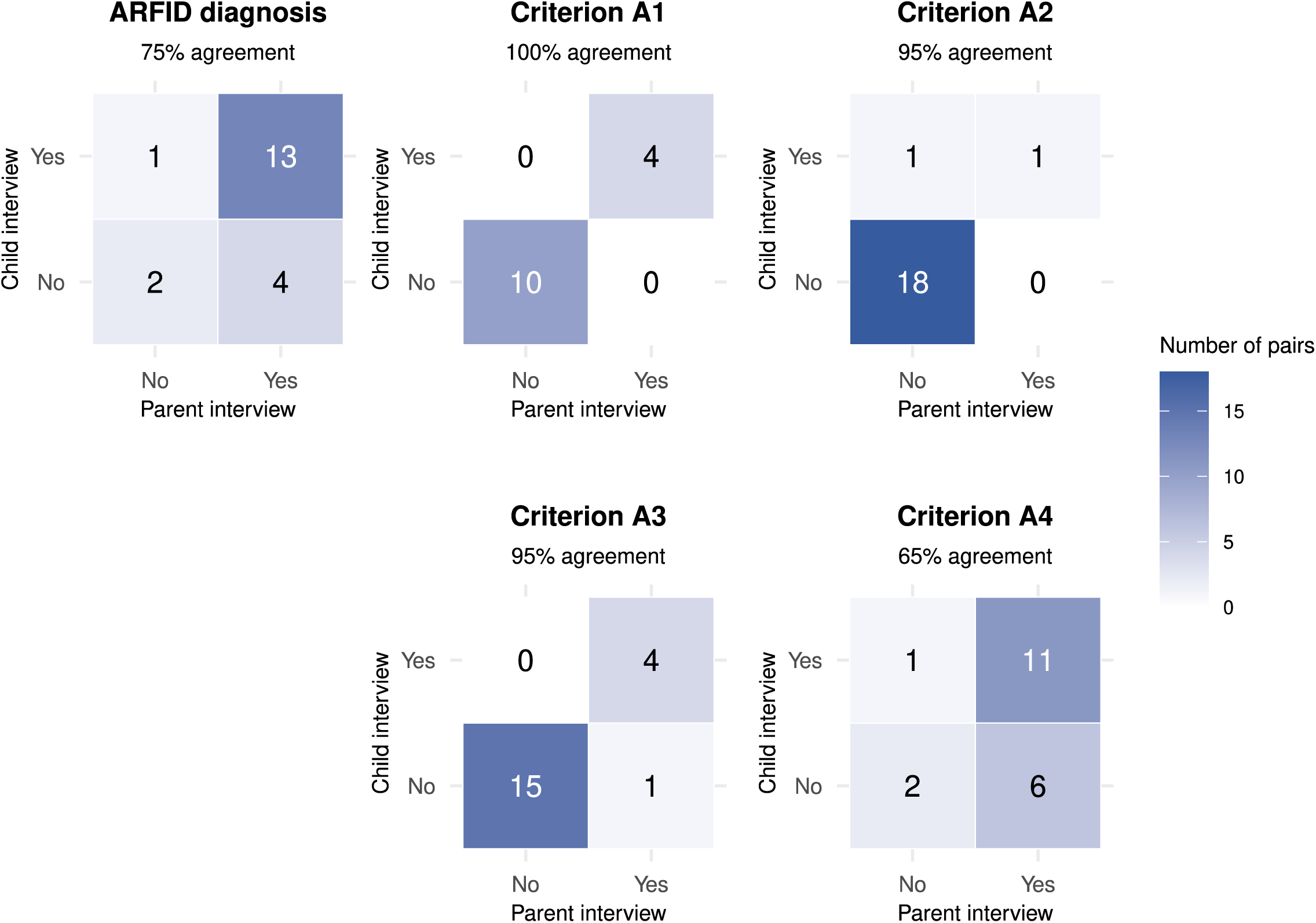
Parent-child agreement for ARFID diagnosis and Criterion A consequences. Columns represent parent interview classifications and rows represent child interview classifications; cells show the number of pairs. ARFID = avoidant/restrictive food intake disorder; Criterion A1 = weight loss or faltering growth; Criterion A2 = nutritional deficiency; Criterion A3 = dependence on enteral feeding or oral nutritional supplements; Criterion A4 = psychosocial impairment. For analysis of Criterion A1, six pairs were removed due to anthropometric information only being available from one informant.

**Table 4.** Parent-child agreement for ARFID diagnosis and Criterion A consequences (categorical ratings).

|  |  |  | **Agreement** |  |  | **Systematic differences** |  |
|---|---|---|---|---|---|---|---|
| **Outcome** | **Parent Criterion met, %** | **Child Criterion met, %** | **Agreement, %** | **Kappa** | **p** | **McNemar’s χ<sup>2</sup>** | **p** |
| ARFID diagnosis | 85.0 | 70.0 | 75.0 | 0.31 | .133 | 0.80 | .371 |
| Criterion A1 | 28.6 | 28.6 | 100.0 | 1.00 | <.001 | NA | NA |
| Criterion A2 | 5.0 | 10.0 | 95.0 | 0.64 | .002 | 0.00 | 1.000 |
| Criterion A3 | 25.0 | 20.0 | 95.0 | 0.86 | <.001 | 0.00 | 1.000 |
| Criterion A4 | 85.0 | 60.0 | 65.0 | 0.19 | .306 | 2.29 | .131 |

**Note.** Criterion A1 = Weight loss or faltering growth; Criterion A2 = Nutritional deficiency; Criterion A3 = Dependence on enteral feeding or oral nutritional supplements; Criterion A4 = Psychosocial impairment; ARFID = Avoidant/restrictive food intake disorder; NA = Not applicable. McNemar’s test compares the two directions of disagreement: parent criterion met/child criterion not met versus parent criterion not met/child criterion met. A significant result indicates that disagreement occurs more often in one direction than would be expected by chance. For analysis of Criterion A1, six pairs were removed due to anthropometric information only being available from one informant. McNemar’s test was not calculated for Criterion A1 because agreement was 100%.

Agreement was higher for the more readily observable consequences. Criterion A1 was met in four parent and four child interviews, with perfect classification agreement (100%; κ = 1.00). Criterion A2 was met in one parent and two child interviews, yielding 95% agreement (κ = 0.64). Criterion A3 was met in five parent and four child interviews, with 95% agreement (κ = 0.86). In contrast, Criterion A4 was met in 17 parent interviews and 12 child interviews, resulting in 65% agreement (κ = 0.19)—the lowest of the four consequences. McNemar’s tests were not statistically significant for Criteria A2, A3, or A4.

### Item-level agreement

Across the 54 PARDI items, parents gave higher mean ratings on 35 items (64.8%), children gave higher ratings on 15 (27.8%), and ratings were equal on four (7.4%; Figure 4). The overall mean parent-minus-child difference on item ratings (scale 0–6) was 0.24 (SD = 0.49, range: -0.89–1.45) and differed significantly from zero (t(53) = 3.57, p < .001), indicating an overall tendency toward higher parental ratings across items. Although parental ratings were generally higher, the direction of informant differences varied across individual PARDI items. The largest differences in the direction of higher parental ratings were observed for difficulty meeting calorie needs (item 33), impact of eating difficulties on family relationships and everyday family life (item 40), difficulties at home involving arguments or conflict with family members (item 41), stressful mealtimes (item 42), and needing prompting or reminding to eat (item 64). The largest differences in the direction of higher child ratings were observed for restricted intake of vegetables, dairy products, and carbohydrates (items 29b, 29d, and 29e), recent loss of interest in food (item 31), reliance on distraction during meals (item 44), discomfort with messy or sticky foods (item 50), difficulty recognizing hunger cues (item 60), and restriction due to fear of abdominal pain (item 74b).

**Figure 4.**
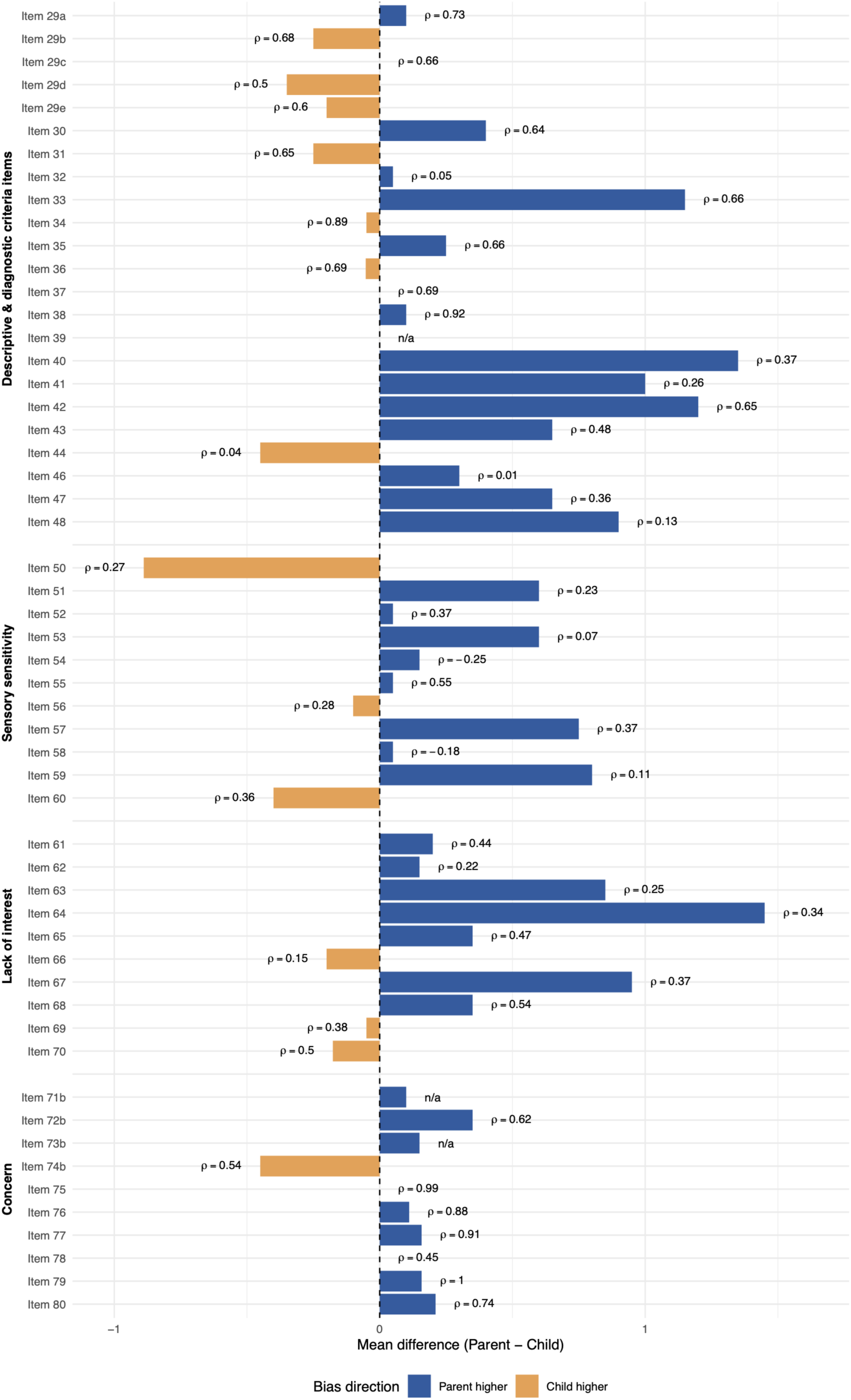
Item-level cross-informant differences and rank-order correspondence. Bars show mean parent-minus-child ratings; positive values indicate higher parent ratings and negative values indicate higher child ratings. Labels report Spearman’s rho (ρ), the correlation between parent and child ratings. Spearman’s rho is undefined (’n/a’) if one or both raters had no variability (item 39: tube feeding, item 71b: concern about choking, item 73b: concern that food may be harmful). ARFID = Avoidant/restrictive food intake disorder; PARDI = Pica, ARFID, and Rumination Disorder Interview.

Rank-order correspondence between parent and child ratings varied substantially, with Spearman’s ρ ranging from −0.25 for strong temperature preferences (item 54) to 1.00 for avoidance related to feared consequences such as vomiting, choking, allergic reactions, or pain (item 79), with a median of 0.47. High correlations were observed for recent weight stagnation (item 34; ρ = .89) and reliance on oral nutritional or high-energy drinks (item 38; ρ = .92), as well as fears of vomiting (item 75; ρ = .99), choking (item 76; ρ = .88), and allergic reactions (item 77; ρ = .91). In contrast, correspondence was weaker for trying new foods (item 32; ρ = .05), distraction during meals (item 44; ρ = .04), encouragement to eat (item 46; ρ = .01), food-color-based avoidance (item 58; ρ = -.18), and eating less during positive emotional states (item 66; ρ = .15). Taken together, findings indicate that the degree of parent–child correspondence depends strongly on the specific symptom being assessed.

## Discussion

This study evaluated parent–child agreement on the PARDI in children aged 11–14 years from the ARIES cohort. Diagnostic classifications were often concordant, while agreement across individual criteria ranged from very high for several nutritional and physical consequences to substantially lower for psychosocial impairment. Among the ARFID dimensions, agreement was strongest for concern about aversive consequences and weakest for sensory sensitivity; parents also rated overall severity significantly higher than children. At the item level, parents generally gave higher ratings, although both the direction of rating differences and rank-order correspondence varied across individual items. Together, these findings indicate that parent and child PARDI interviews capture overlapping but non-equivalent aspects of ARFID.

Despite the overall diagnostic concordance, five of the 20 (25.0%) parent–child pairs were not concordant. Four were classified as ARFID based on the parent interview but not the child interview, while the reverse occurred in one pair. These discordant cases indicate that differences in cross-informant perspectives can affect diagnostic outcome.

Looking beyond the overall diagnosis, agreement differed markedly across the individual diagnostic criteria. Parent–child agreement was high for weight or growth, nutritional deficiency, and dependence on nutritional supplementation (Criteria A1-A3), but substantially lower for psychosocial impairment (Criterion A4). This pattern may reflect differences in how readily these consequences can be observed or documented. Information on growth, nutritional deficiency, and nutritional support may be available from measurements, healthcare contacts, or established treatment routines, whereas psychosocial impairment is more dependent on context and perspective. Parents may be more aware of how the child’s eating difficulties affect shared meals, eating outside the home, or family routines, whereas children may be more aware of anxiety or embarrassment that parents do not experience firsthand. Children may also minimize the psychosocial impact/consequences of their eating difficulties, including difficulties in interpersonal or social functioning, describing the consequences as less severe or disruptive, which could contribute to disagreement when parents report greater impairment. The lower agreement for A4 may therefore reflect differences in how psychosocial impairment is experienced, observed, or reported across family, mealtime, and social contexts.

Agreement also varied across the three ARFID dimensions. Agreement for concern about aversive consequences was excellent, contrasting with poorer agreement for sensory sensitivity and lack of interest. The excellent agreement for concern should, however, be interpreted with care. Concern scores were low for most participants, and clustering near zero can produce apparent concordance when both informants consistently report absence of the construct. When concern was present, however, items describing specific feared outcomes also showed very high rank-order correspondence. Fear-related episodes may be salient, discrete, and readily communicated within families, whereas sensory dislike and diminished appetite may be persistent but less easily observed or verbalized.

One clear difference between informants emerged for overall ARFID severity, which parents rated significantly higher than children. Although parent means were also numerically higher across ARFID dimensions, none of these differences was statistically significant. The higher parent-rated severity may partly reflect aspects of the eating difficulties that are particularly apparent to parents in everyday life. Several of the more pronounced parent-higher item differences concerned difficulty meeting nutritional needs, stressful mealtimes, and the impact of eating difficulties on home and family life.

Item-level analyses provided a more detailed picture of these informant differences. Although parents reported higher mean ratings on most items, children reported higher ratings on some items, including restriction of particular food groups, dropping previously liked foods, reliance on distraction during meals, discomfort with messy or sticky foods, difficulty recognizing hunger cues, and restriction because of fear of abdominal pain. For some of these items, such discrepancies may become even more likely as children move into adolescence and gain greater independence around eating. More meals may then occur outside the family home, reducing opportunities for direct parental observation. Rank-order correspondence also varied substantially across items. Parent and child ratings corresponded closely for recent lack of expected weight gain, reliance on oral nutritional or high-energy drinks, and specific fears of vomiting, choking, and allergic reactions, whereas correspondence was weak for willingness to try new foods, distraction during meals, encouragement to eat, food-color-based avoidance, and eating less during positive emotional states. Together, these findings indicate that both the direction of informant differences and the degree of parent–child correspondence vary considerably across individual PARDI items.

The present findings are broadly consistent with previous cross-informant research in ARFID. Using the NIAS, Carmody et al. (9) found that caregivers generally reported greater ARFID symptom severity than their children, resembling the overall parent-higher pattern observed in the present study. Barnhart et al. (12) found a similar but non-significant pattern across the NIAS subscales. Validation of the EDE–ARFID module similarly showed that parent–child agreement varied across individual diagnostic criteria, although overall diagnostic agreement was substantially higher than in the present study (75%): 94% for the child–parent comparison and 96% for the adolescent–parent comparison (10). The reasons for this difference are not entirely clear. One possible explanation lies in sample composition. Their cohort included non-treatment-seeking controls, none of whom met criteria for current ARFID, whereas our sample consisted entirely of screen-positive participants. This may have increased the likelihood of concordant negative classifications in their sample, while different parent and child accounts in our sample had greater potential to result in different diagnostic classifications. Taken together, these findings reinforce the central point that even though child and parent assessments of ARFID overlap, considering both perspectives can uncover meaningful differences that either assessment alone may miss.

### Strengths and limitations

The current study has several strengths. It addresses the previously unexamined cross-informant agreement across diagnostic, dimensional, and item-level outcomes of the PARDI. Parent and child interviews were conducted separately by different interviewers, reducing direct carryover of information between assessments. Recurring consensus meetings supported standardized scoring, and double ratings showed moderate to excellent interviewer reliability across outcomes.

The principal limitation is the small sample size, which reduced statistical power and produced wide confidence intervals around several agreement estimates. In particular, the study had limited power to detect systematic directional differences in discordant diagnostic classifications. The relatively small sample size also prevented us from investigating potential moderators of parent–child agreement such as child sex, autistic traits, or anxiety traits. Furthermore, the sample was restricted to children aged 11–14 years who had screened positive for ARFID in a large-scale epidemiological study, limiting generalizability to other age groups and populations, such as treatment-seeking or clinically diagnosed individuals. Importantly, screening status in the ARIES study was determined using parent-report questionnaires, which may have preferentially selected for parent-visible forms of ARFID, potentially influencing the parent–child agreement patterns observed in this study.

Anthropometric information was reported rather than directly measured, which may have reduced the precision of the information used to assess Criterion A1. Further, the observed agreement patterns may differ in larger and more diverse samples, for example, among younger children (<11 years) or adolescents >14 years, clinically diagnosed or treatment-seeking individuals, screen-negative participants, or samples with a more balanced representation of mothers and fathers as parent informants. These findings should therefore be regarded as preliminary estimates rather than fixed properties of the PARDI.

### Clinical and research implications

The findings support a cross-informant approach when the PARDI is used with children. When child and parent accounts differ, the PARDI provides an opportunity to clarify how the child’s functioning is affected across different contexts and what each informant has observed. If only one interview is possible, clinicians should be aware that ratings may vary depending on the informant, particularly for severity and psychosocial impairment.

Importantly, the present study compared child and parent PARDI interviews rather than treating either reporter as the reference against which the other was judged. For some aspects of ARFID, one informant may have access to more accurate or readily verifiable information, whereas for others, differences may reflect genuinely different experiences or observations. Further, there was no separate clinical assessment integrating both accounts with anthropometric, medical, and functional information. Higher parental ratings should therefore not be interpreted as parent over-reporting, nor lower child ratings as child under-reporting. Instead, these discrepancies may identify areas that require further clarification during assessment. This may be particularly relevant for psychosocial impairment, where parent–child agreement was lowest and where considering both perspectives may therefore be especially informative during assessment.

Larger studies should estimate agreement across age groups, ARFID dimensions, sexes, cultural contexts, and clinical settings. They should include multiple caregivers where possible, document which caregiver is most involved in meals, and incorporate direct anthropometric measurement, relevant medical records, and clinician consensus. For epidemiological and treatment research, the choice of informant may also affect case definition, baseline severity, and apparent change over time. Studies should prespecify whether diagnosis is based on parent report, child report, or clinical integration, and should use the same approach at follow-up. Reporting both informant-specific and integrated results would make findings more comparable and help determine whether discrepancies themselves carry prognostic information.

## Conclusion

Parent and child PARDI interviews showed meaningful but incomplete agreement in children screening positive for ARFID on parent-reported questionnaires. Concordance was strongest for more readily observable consequences and concern about aversive outcomes, whereas sensory sensitivity, lack of interest in food or eating, overall diagnosis, and especially psychosocial impairment were more informant-dependent. Parents generally reported greater overall severity, whereas children endorsed some specific eating experiences more strongly, such as difficulty recognizing hunger cues, discomfort with messy or sticky foods, and fear of abdominal pain. These findings support integrating multiple perspectives in pediatric ARFID assessment and provide a basis for larger studies of the PARDI’s cross-informant validity.

## Author Contribution Statement

**Dahlbäck:** investigation, data curation; visualization; writing – original draft preparation.

**Moberg:** visualization; writing – original draft preparation.

**Friskson:** investigation; data curation; writing – review and editing.

**Myrberg:** investigation, data curation; writing – review and editing.

**Hog**: conceptualization; methodology; resources; investigation; data curation; writing – review and editing.

**Micali**: validation; writing – review and editing.

**Bulik**: conceptualization; funding acquisition; writing – review and editing.

**Dinkler**: conceptualization; funding acquisition; methodology; resources; investigation; data curation; formal analysis; visualization; project administration; supervision; writing – review and editing.

## Data Availability

Data and code that support the findings of this study are available from the corresponding author upon reasonable request.

## Ethical Statement

This study was approved by the Swedish Ethical Review Authority (no. 2023-04638-0, 2025-01356-02).

## Acknowledgements

We would like to express our sincere gratitude to the ARFID Initiative Sweden (ARIES) participants, the ARIES Parent Advisory Council, and research nurse Martina Wennberg.

## Disclosure statement about AI use tools

ChatGPT 5.2 was used to write and debug code for data analysis and for proofreading and improving grammar and writing clarity.

## Involvement of persons with lived experiences

The ARFID Initiative Sweden (ARIES) engages a Parent Advisory Council comprising eight parents/guardians of children and adolescents with ARFID to ensure that the research is feasible, relevant, and beneficial to the community. Prior to the main study launch, the Parent Advisory Council consulted with researchers on the selection of questionnaires, completed a beta version of the full study, and provided feedback on all study procedures. The Parent Advisory Council was not involved in the study design, execution, or write-up of the study described here.

